# Health Needs Assessment: Development and Validation of a Simplified Risk Probability Scale (SRPS) for Mental Health Screening in COVID-19 Frontline Rescue Teams

**DOI:** 10.1101/2025.02.09.25321964

**Authors:** Yan Bo, Yifei Chen, Fei Zhao

## Abstract

Frontline rescue workers during the COVID-19 pandemic faced heightened psychological risks, yet existing tools like PHQ-9 and GAD-7 lack crisis-specific relevance and brevity. This study aimed to develop and validate a simplified risk probability scale (SRPS) tailored for rapid self-assessment of depression and anxiety in frontline rescue teams. A cross-sectional survey of 273 frontline workers in Lanzhou, China, was conducted using snowball sampling. Participants completed questionnaires integrating socio-economic, occupational, and psychological metrics, analyzed via univariate, multivariate, and ROC curve analyses. Key findings revealed middle-income earners (3000–4000 CNY/month) faced elevated depression risk [OR=3.666, 95%CI (1.085-12.385), p=0.036], while work stress intolerance strongly predicted anxiety [OR=14.258, 95%CI (4.213-58.983), p<0.001]. The SRPS demonstrated moderate predictive accuracy (depression AUC=0.572, sensitivity=58.2%, specificity=53.7%; anxiety AUC=0.662, sensitivity=72.5%, specificity=64.8%)] but prioritized brevity (10 items) and contextual relevance over diagnostic precision. The tool’s integration into mobile health platforms offers real-time screening potential, enabling targeted interventions for high-risk groups. This study highlights the necessity of context-specific mental health tools in crisis settings and provides a foundation for scalable, dynamic risk assessment in future public health emergencies.

## 1 Introduction

The COVID-19 pandemic has profoundly disrupted global daily life since its emergence in December 2019. Characterized by rapid progression to pulmonary fibrosis and hypoxia-related mortality ^1^, the virus’s high mutation rate has rendered conventional antiviral therapies largely ineffective, necessitating stringent public health measures such as restricted outdoor activities for self-protection ^1^. While China initially pursued herd immunity strategies to mitigate pandemic spread ^2^, the Hong Kong government implemented a containment approach involving non-medical frontline personnel, including security personnel and volunteers, to curb viral transmission ^3^.

Frontline rescue workers constitute a high-risk population for psychological distress due to prolonged exposure to occupational stressors, including heavy workloads, infection risks, and social isolation from family members ^4,5^. Studies indicate anxiety and depression prevalence rates of 19.8% and 27.7%, respectively, among Hong Kong’s anti-epidemic teams ^6^. Key risk factors encompass COVID-19 infection, emotional isolation, familial separation, logistical constraints, and age-related vulnerabilities ^4,7–9^. However, existing assessment tools like PHQ-9 and GAD-7 exhibit critical limitations in crisis contexts: their generic design lacks specificity for rescue operations, and their administration time (10–15 minutes) proves impractical during high-pressure missions ^10^.

This gap underscores the urgent need for context-specific psychological risk assessment tools. A validated predictive model incorporating pandemic-specific variables—such as income instability, rotational work systems, and occupational stressors—could enable rapid self-assessment through simplified scoring protocols ^10^. Such innovation requires interdisciplinary collaboration between mental health professionals and rescue organizations to ensure ecological validity and operational feasibility.

The present study aims to address these challenges by developing a simplified risk probability scale (SRPS) tailored for frontline anti-epidemic personnel. Distinct from conventional instruments, the SRPS prioritizes brevity (10 items) and contextual relevance, integrating socio-economic and occupational predictors unique to pandemic rescue operations. By enabling real-time mental health screening, this tool holds potential for integration into mobile health platforms, thereby enhancing early intervention capabilities during public health emergencies.

## 2 Results

### 2.1 Participant characteristics

During the questionnaire survey from 1st October to 1st November 2022, a total of 293 participants were surveyed in the SRSP study, of which 20 participants gave up partially answering the questionnaires, with a return rate of 100% (**Figure 1**).

**Figure 1.**
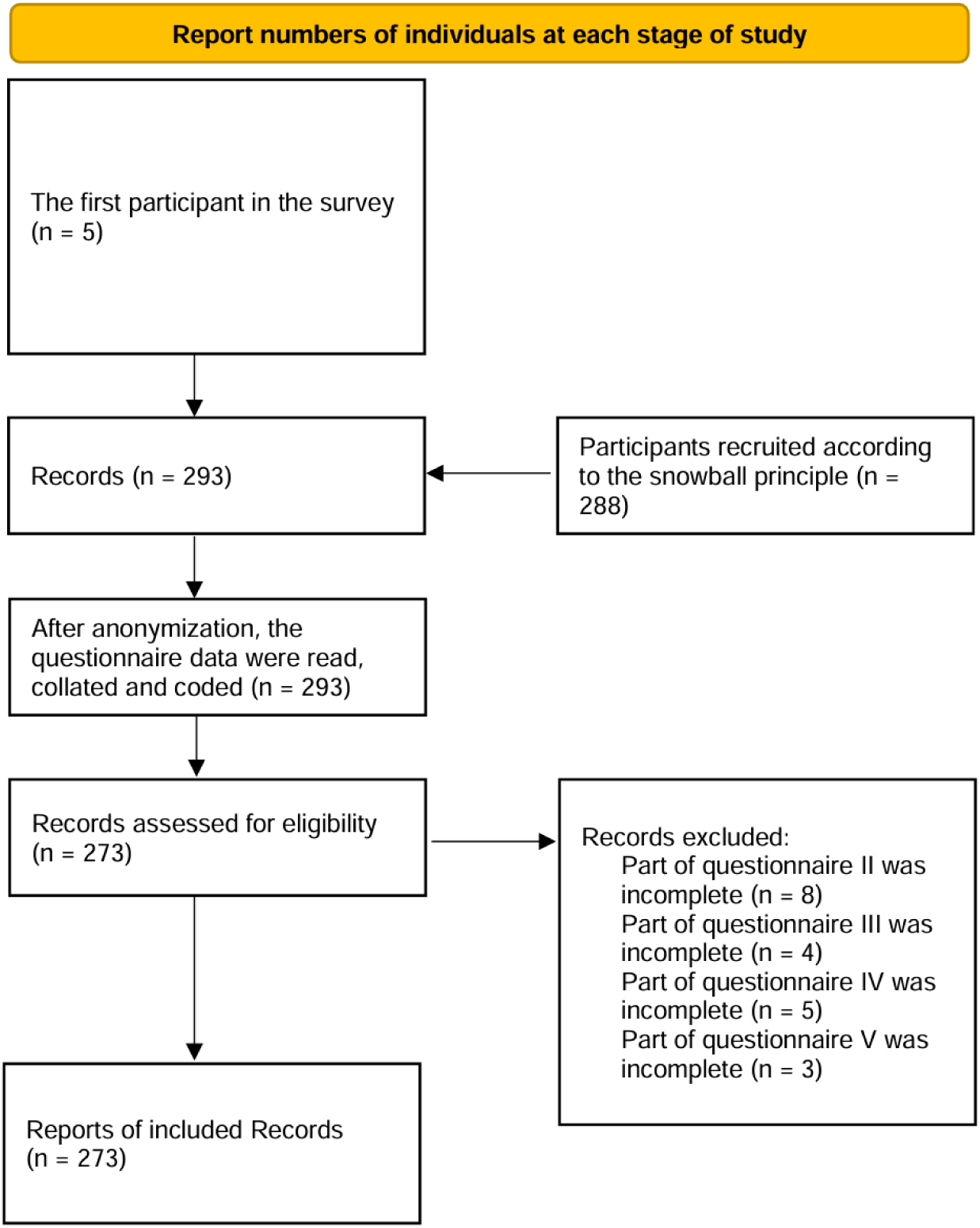
Flow diagram. This flow chart described the details of participants from inclusion to final exclusion during the questionnaire. Finally, only the completed questionnaires would be included in the report, and those incomplete questionnaires would be excluded.

The study enrolled 273 frontline rescue workers (64.8% female; mean age 32.4±8.7 years) with complete questionnaire responses. Depression and anxiety symptom prevalence rates were 34.1% (93/273) and 35.9% (98/273), respectively. Key demographic characteristics are summarized in Table 1.

**Table 1.**
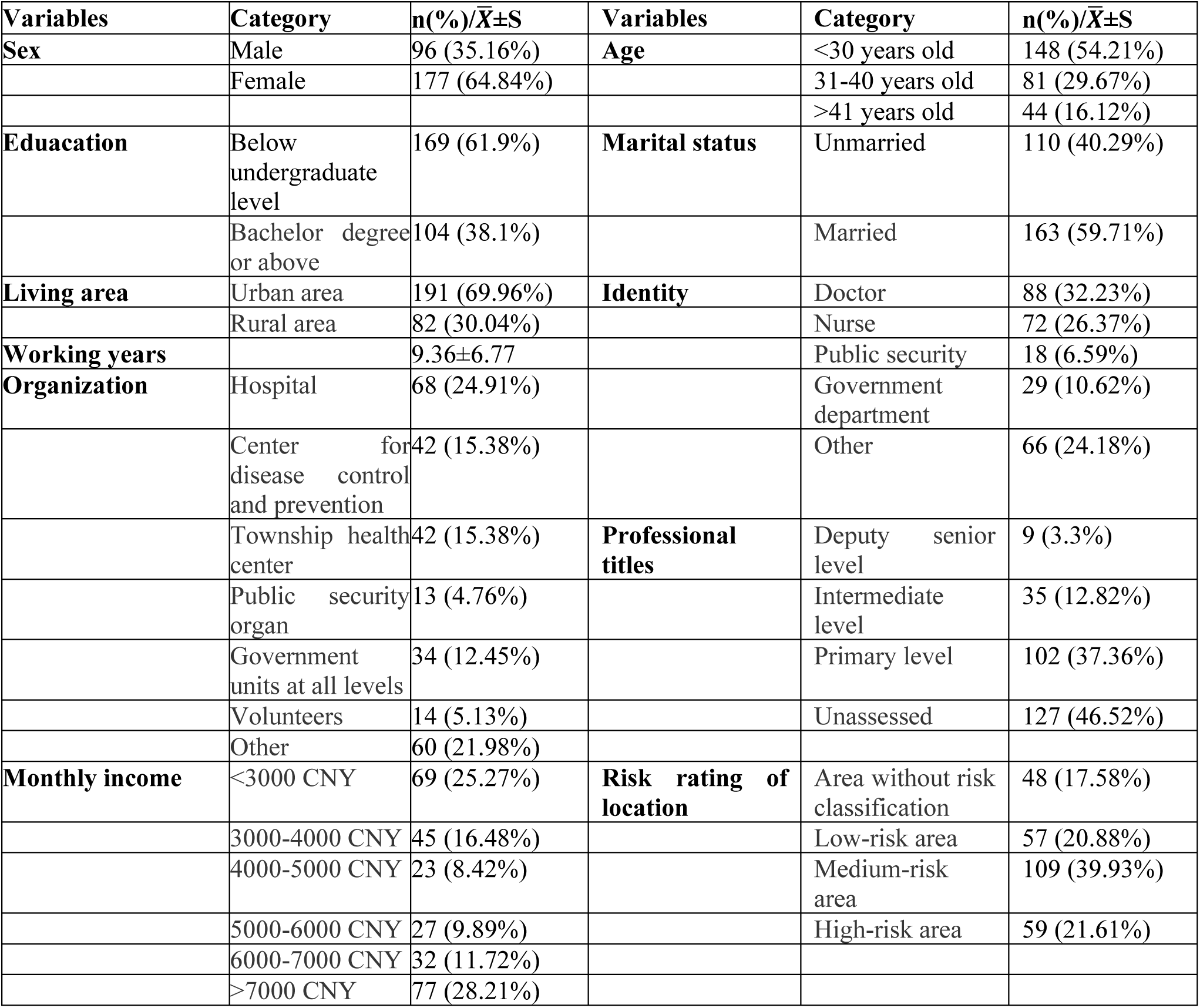
Demographics. This table showed the comprehensive demographic information of the 273 frontline rescue workers.

| Variables | Category | n(%)/$\bar{X}\pm S$ | Variables | Category | n(%)/$\bar{X}\pm S$ |
| --- | --- | --- | --- | --- | --- |
| **Sex** | Male | 96 (35.16%) | **Age** | <30 years old | 148 (54.21%) |
|  | Female | 177 (64.84%) |  | 31-40 years old | 81 (29.67%) |
|  |  |  |  | >41 years old | 44 (16.12%) |
| **Education** | Below undergraduate level | 169 (61.9%) | **Marital status** | Unmarried | 110 (40.29%) |
|  | Bachelor degree or above | 104 (38.1%) |  | Married | 163 (59.71%) |
| **Living area** | Urban area | 191 (69.96%) | **Identity** | Doctor | 88 (32.23%) |
|  | Rural area | 82 (30.04%) |  | Nurse | 72 (26.37%) |
| **Working years** | | 9.36±6.77 | | Public security | 18 (6.59%) |
| **Organization** | Hospital | 68 (24.91%) |  | Government department | 29 (10.62%) |
|  | Center for disease control and prevention | 42 (15.38%) |  | Other | 66 (24.18%) |
|  | Township health center | 42 (15.38%) | **Professional titles** | Deputy senior level | 9 (3.3%) |
|  | Public security organ | 13 (4.76%) |  | Intermediate level | 35 (12.82%) |
|  | Government units at all levels | 34 (12.45%) |  | Primary level | 102 (37.36%) |
|  | Volunteers | 14 (5.13%) |  | Unassessed | 127 (46.52%) |
|  | Other | 60 (21.98%) |  |  |  |
| **Monthly income** | <3000 CNY | 69 (25.27%) | **Risk rating of location** | Area without risk classification | 48 (17.58%) |
|  | 3000-4000 CNY | 45 (16.48%) |  | Low-risk area | 57 (20.88%) |
|  | 4000-5000 CNY | 23 (8.42%) |  | Medium-risk area | 109 (39.93%) |
|  | 5000-6000 CNY | 27 (9.89%) |  | High-risk area | 59 (21.61%) |
|  | 6000-7000 CNY | 32 (11.72%) |  |  |  |
|  | >7000 CNY | 77 (28.21%) |  |  |  |

### 2.2 Univariate analysis

Questionnaire data collected during the SRPS study revealed that participants’ monthly income was a significant single factor influencing the presence of depressive symptoms (χ2=11.63, P<0.05). Interestingly, the number of depressive symptoms examined was the lowest (n=6) for participants with medium monthly income as a percentage of the overall level (monthly income between 3000 and 4000), while the number of depressive symptoms examined was the highest (n=32) for participants with the highest monthly income as a percentage of the overall level (monthly income more than 7000). From the perspective of anxiety, participants’ age group (χ2=16.35, P<0.05), marital status (χ2=11.86, P<0.05), and risk level of the area of residence (χ2=8.94, P<0.05) were the significant monofactors influencing the presence of anxiety. Of these single factors, participants living in areas with a high risk rating had the highest number of detected anxiety (n=31), while the lowest detection of anxiety was not in unclassified risk areas (n=20) but rather in low risk areas (n=18). The unclassified risk area rating represents an area where patients infected with COVID-19 do not yet exist, which may be related to the distribution of social rescuers’ residences. Participants who were married had a higher number of detected anxiety compared to those who were unmarried (n=101), which may be related to the family responsibilities that come with marriage. Participants who were no older than 30 years old had the highest number of detected anxiety (n=62), while those who were no younger than 41 years old had the lowest number of detected anxiety (n=29), which seems to imply that participants older than 41 years old are more tolerant of anxiety (**Table 2** and **Table 3**).

**Table 2.** Chi-square analysis of depression symptoms and anxiety symptoms in the context of demographic information. This table showed the outcomes of univariate analyses regarding depression symptoms and anxiety symptoms among frontline rescue workers, with a focus on the perspective of demographic information.

| Variables | Category | Depression symptoms<br>n(%) | | $\chi^2$ | P | Anxiety symptoms<br>n(%) | | $\chi^2$ | P |
| --- | --- | --- | --- | --- | --- | --- | --- | --- | --- |
|  |  | Yes | No |  |  | Yes | No |  |  |
| Age |  |  |  | 2.79 | 0.25 |  |  | 16.35 | <0.05* |
|  | <30 years old | 44 (16.12%) | 104 (38.1%) |  |  | 86 (31.5%) | 62 (22.71%) |  |  |
|  | 31-40 years old | 31 (11.36%) | 50 (18.32%) |  |  | 27 (9.89%) | 54 (19.78%) |  |  |
|  | >41 years old | 18 (6.59%) | 26 (9.52%) |  |  | 15 (5.49%) | 29 (10.62%) |  |  |
| Marital status |  |  |  | 0.65 | 0.42 |  |  | 11.86 | <0.05* |
|  | Unmarried | 54 (19.78%) | 115 (42.12%) |  |  | 66 (24.18%) | 44 (16.12%) |  |  |
|  | Married | 39 (14.29%) | 65 (23.81%) |  |  | 62 (22.71%) | 101 (37%) |  |  |
| Monthly income |  |  |  | 11.63 | <0.05* |  |  | 2.09 | 0.84 |
|  | <3000 CNY | 24 (8.79%) | 45 (16.48%) |  |  | 34 (12.45%) | 35 (12.82%) |  |  |
|  | 3000-4000 CNY | 6 (2.2%) | 39 (14.29%) |  |  | 19 (6.96%) | 26 (9.52%) |  |  |
|  | 4000-5000 CNY | 10 (3.66%) | 13 (4.76%) |  |  | 9 (3.3%) | 14 (5.13%) |  |  |
|  | 5000-6000 CNY | 9 (3.3%) | 18 (6.59%) |  |  | 12 (4.4%) | 15 (5.49%) |  |  |
|  | 6000-7000 CNY | 12 (4.4%) | 20 (7.33%) |  |  | 14 (5.13%) | 18 (6.59%) |  |  |
|  | >7000 CNY | 32 (11.72%) | 45 (16.48%) |  |  | 40 (14.65%) | 37 (13.55%) |  |  |
| Risk rating of location |  |  |  | 2.26 | 0.52 |  |  | 8.94 | <0.05* |
|  | Area without risk classification | 16 (5.86%) | 32 (11.72%) |  |  | 20 (7.33%) | 28 (10.26%) |  |  |
|  | Low-risk area | 19 (6.96%) | 38 (13.92%) |  |  | 18 (6.59%) | 39 (14.29%) |  |  |
|  | Medium-risk area | 42 (15.38%) | 67 (24.54%) |  |  | 59 (21.61%) | 50 (18.32%) |  |  |
|  | High-risk area | 16 (5.86%) | 43 (15.75%) |  |  | 31 (11.36%) | 28 (10.26%) |  |  |

**Table 3.**
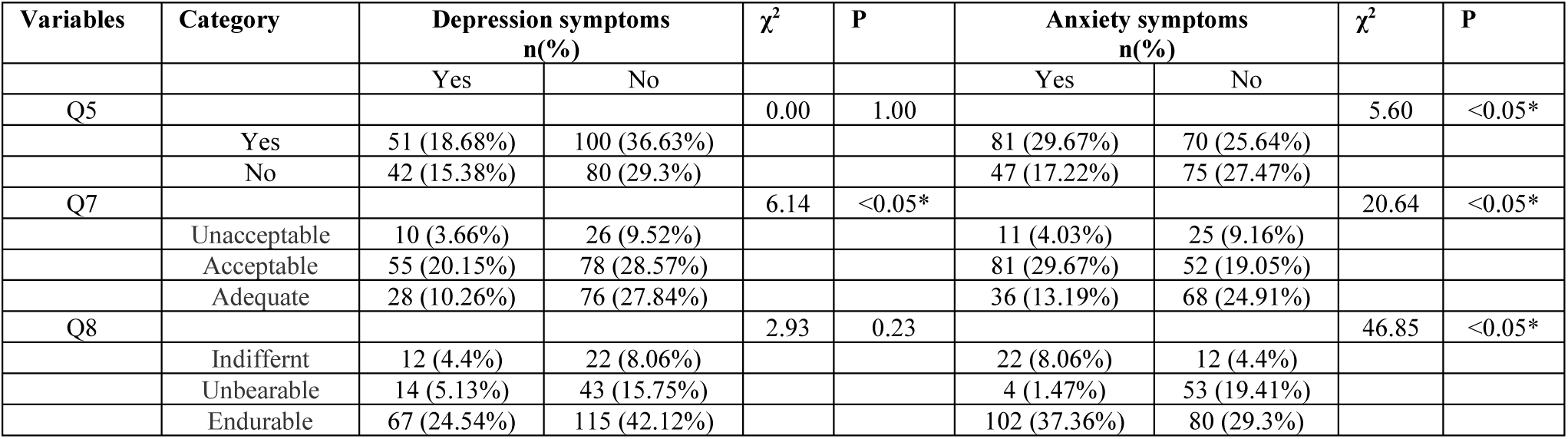
Chi-square analysis of participants’ depression symptoms and anxiety symptoms in the context of COVID-19. This table showed the results of univariate analyses of depression and anxiety symptoms among frontline rescue workers, with a specific focus on the impact of the COVID-19 pandemic. Additionally, the questions in the questionnaire included: Q5 “Are there any confirmed cases with you or those around you?”; Q7 “What do you think of the shift system?”; Q8 “How do you perceive the work pressure?”

| Variables | Category | Depression symptoms<br>n(%) | | $\chi^2$ | P | Anxiety symptoms<br>n(%) | | $\chi^2$ | P |
| --- | --- | --- | --- | --- | --- | --- | --- | --- | --- |
|  |  | Yes | No |  |  | Yes | No |  |  |
| Q5 |  |  |  | 0.00 | 1.00 |  |  | 5.60 | <0.05* |
|  | Yes | 51 (18.68%) | 100 (36.63%) |  |  | 81 (29.67%) | 70 (25.64%) |  |  |
|  | No | 42 (15.38%) | 80 (29.3%) |  |  | 47 (17.22%) | 75 (27.47%) |  |  |
| Q7 |  |  |  | 6.14 | <0.05* |  |  | 20.64 | <0.05* |
|  | Unacceptable | 10 (3.66%) | 26 (9.52%) |  |  | 11 (4.03%) | 25 (9.16%) |  |  |
|  | Acceptable | 55 (20.15%) | 78 (28.57%) |  |  | 81 (29.67%) | 52 (19.05%) |  |  |
|  | Adequate | 28 (10.26%) | 76 (27.84%) |  |  | 36 (13.19%) | 68 (24.91%) |  |  |
| Q8 |  |  |  | 2.93 | 0.23 |  |  | 46.85 | <0.05* |
|  | Indifferent | 12 (4.4%) | 22 (8.06%) |  |  | 22 (8.06%) | 12 (4.4%) |  |  |
|  | Unbearable | 14 (5.13%) | 43 (15.75%) |  |  | 4 (1.47%) | 53 (19.41%) |  |  |
|  | Endurable | 67 (24.54%) | 115 (42.12%) |  |  | 102 (37.36%) | 80 (29.3%) |  |  |

#### Depression Symptoms

Monthly income demonstrated significant associations with depression (χ²=11.63, p<0.05). Paradoxically, the lowest depression prevalence was observed in the 3000-4000 CNY group (13.3%, 6/45), while the highest rates occurred in the >7000 CNY cohort (41.6%, 32/77). Acceptance of shift rotation systems emerged as another significant predictor (χ²=6.14, p=0.047), with higher depression rates among rotation-accepting workers (41.4%, 55/133).

#### Anxiety Symptoms

Younger age (χ²=16.35, p<0.001), marital status (χ²=11.86, p=0.001), and high-risk residential areas (χ²=8.94, p=0.03) were significantly associated with anxiety. Married participants aged ≤30 years in high-risk zones exhibited the highest anxiety prevalence (52.9%, 31/59). Intolerance to work pressure showed protective effects against anxiety (χ²=46.85, p<0.001), with only 8.2% (12/146) affected. We found by analysing participants ‘responses to questions related to rescue COVID-19 that the attitude maintained by the rescue rotation system faced by participants during COVID-19 was a significant single factor influencing participants’ acquisition of depressive symptoms (χ2=6.14, P<0.05). The number of participants who were able to accept the rotation system who detected depressive symptoms was the highest (n=55), while those who could not tolerate it were the lowest (n=10). This implies that depressive symptoms may arise from being forced to perform social rescue tasks, otherwise social rescuers would not be faced with the rotation system. From the perspective of anxiety, the diagnosis of COVID-19 infection by the participant or the participant’s friend (χ2=5.60, P<0.05), the attitude maintained by the rescue rotation system faced by the participant during the COVID-19 period (χ2=20.64, P<0.05*), and the participant’s attitude towards facing the stress of work during the COVID-19 period (χ2=46.85, P <0.05) were significant single factors influencing participants’ acquisition of anxiety. Here we found that the rotation system was a significant single factor common to both anxious and depressive symptomss, and that the more symptoms -inducing attitudes of both were surprisingly consistent. This may indicate that participants were annoyed by taking on tasks that were not part of their job description. A diagnosis of COVID-19 infection in a participant or a participant’s friend was more likely to cause participants to experience anxiety (n=81). Interestingly, participants who could not tolerate work stress during COVID-19 were the least likely to experience anxiety (n=12), and we hypothesise that these work stresses are eventually transferred to social rescuers who either do not know how to reject work stresses or are happy to indefinitely crush their egos (**Table 3**).

### 2.3 Multivariate analysis

The AUC for the depressive symptoms model was 347.13 with a pseudo R^2^ of 0.05. We found in the multifactorial analysis that participants’ monthly income of CNY3,000 to CNY4,000 was a risk factor for participants to experience depressive symptoms [odds ratio (OR) = 3.666 (1.085-12.385)] (**Table 4**).

**Table 4.** Logistic regression analysis of influence on depression symptoms. This table showed the outcomes of the multifactorial analysis of factors influencing depression symptoms among frontline rescue workers. Additionally, the questions in the questionnaire include: Q7 “What do you think of the shift system?” In this analysis, multiple essential statistical parameters were presented: OR: Odds ratio; 95%CI: 95% Confidence interval; P: P-value; SE: Standard error; β: Beta coefficient; CNY: Chinese Yuan.

| Variables | Category | $\beta$ | SE | P | OR | 95%CI |
| --- | --- | --- | --- | --- | --- | --- |
| Monthly income | 3000-4000 CNY | 1.299 | 0.621 | 0.036* | 3.666 | 1.085-12.385 |
|  | 4000-5000 CNY | -0.306 | 0.702 | 0.663 | 0.736 | 0.186-2.914 |
|  | 5000-6000 CNY | 0.341 | 0.633 | 0.59 | 1.406 | 0.406-4.867 |
|  | 6000-7000 CNY | 0.022 | 0.559 | 0.968 | 1.023 | 0.342-3.058 |
|  | >7000 CNY | -0.211 | 0.495 | 0.67 | 0.81 | 0.307-2.137 |
| Q7 | Acceptable | -0.598 | 0.534 | 0.263 | 0.55 | 0.193-1.566 |
|  | Adequate | 0.429 | 0.566 | 0.449 | 1.536 | 0.506-4.658 |

Whereas the model AUC for anxiety symptoms was 327.39 with a pseudo R^2^ of 0.20, we found in the multifactorial analysis that participants ‘inability to tolerate work stress during the COVID-19 period was a risk factor for participants’ acquisition of anxious symptoms [OR = 14.258 (4.213-58.983)], which is in contrast to the unifactorial results showing that participants’ inability to tolerate work stress during the COVID-19 period Work stress had the lowest number of anxiety detected (n=12) which differed from the results of the single factor results showing that participants were unable to tolerate the stress of work during COVID-19. It is clear that multifactorial analyses are more likely to provide insight into the intrinsic influences between multiple factors compared to unifactorial analyses (**Table 5**).

**Table 5.**
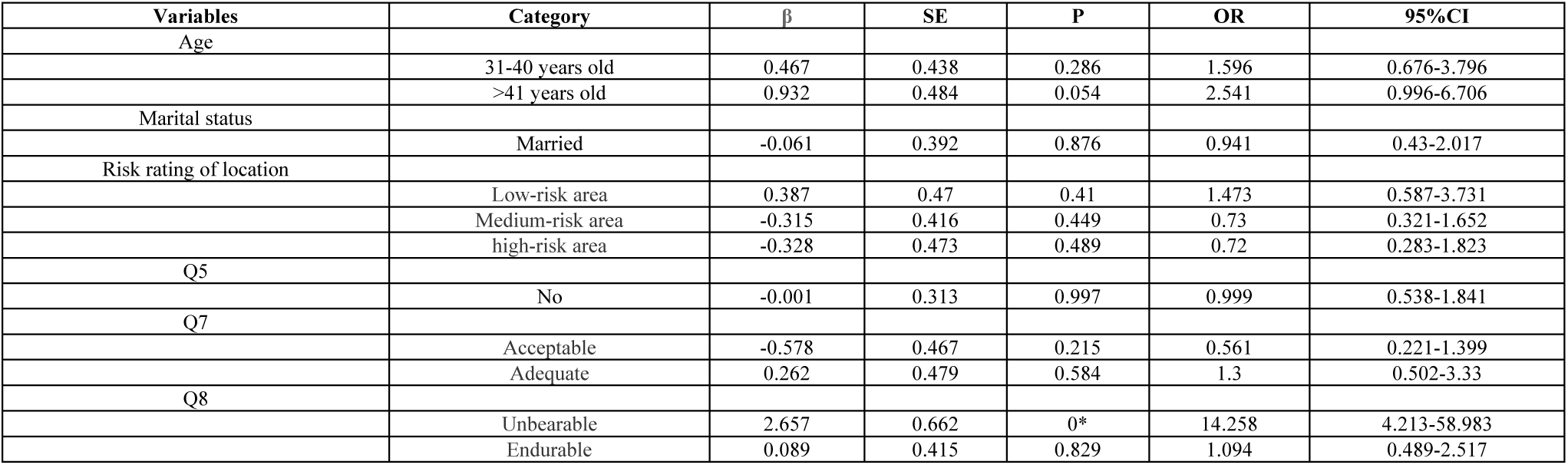
Logistic regression analysis of influencing anxiety symptoms. . This table presented the results of the multifactorial analysis of factors influencing anxiety symptoms among frontline rescue workers. Additionally, the questions in the questionnaire included: Q5 “Are there any confirmed cases with you or those around you?”; Q7 “What do you think of the shift system?”; Q8 “How do you perceive the work pressure?” In this analysis, several key statistical parameters were provided: OR: Odds ratio; 95%CI: 95% Confidence interval; P: P-value; SE: Standard error; β: Beta coefficient.

| Variables | Category | $\beta$ | SE | P | OR | 95%CI |
| --- | --- | --- | --- | --- | --- | --- |
| Age | 31-40 years old | 0.467 | 0.438 | 0.286 | 1.596 | 0.676-3.796 |
|  | >41 years old | 0.932 | 0.484 | 0.054 | 2.541 | 0.996-6.706 |
| Marital status | Married | -0.061 | 0.392 | 0.876 | 0.941 | 0.43-2.017 |
| Risk rating of location | Low-risk area | 0.387 | 0.47 | 0.41 | 1.473 | 0.587-3.731 |
|  | Medium-risk area | -0.315 | 0.416 | 0.449 | 0.73 | 0.321-1.652 |
|  | high-risk area | -0.328 | 0.473 | 0.489 | 0.72 | 0.283-1.823 |
| Q5 |  |  |  |  |  |  |
|  | No | -0.001 | 0.313 | 0.997 | 0.999 | 0.538-1.841 |
| Q7 | Acceptable | -0.578 | 0.467 | 0.215 | 0.561 | 0.221-1.399 |
|  | Adequate | 0.262 | 0.479 | 0.584 | 1.3 | 0.502-3.33 |
| Q8 | Unbearable | 2.657 | 0.662 | 0* | 14.258 | 4.213-58.983 |
|  | Endurable | 0.089 | 0.415 | 0.829 | 1.094 | 0.489-2.517 |

### 2.4 Structural equations

In terms of depression, univariate analyses showed that monthly income was a significant influencing factor, with the lowest rates of depression detection among subjects with monthly incomes between 3,000-4,000 CNY and the highest rates among those with monthly incomes of more than 7,000 CNY; attitudes towards the rescue rotation system also influenced depression, with the highest rates of depression detection among those who were able to accept the rotation system. Univariate analyses showed higher rates of depression among those with incomes below 7,000 CNY, whereas a multifactor model found that middle income (3,000-4,000 CNY) was an independent risk factor (OR=3.67, p=0.036). This difference may stem from confounding variables. This was addressed here by exploring the relationship between confounding variables through structural equations.

The mediating role of job security is manifested in the middle-income group’s lower job security and thus higher risk of depression, possibly due to lower job stability (**Figure 2**). In contrast, the moderating role of family responsibilities is manifested in the fact that married middle-income earners may further amplify depression risk due to the double loading of family financial burdens and work pressures (**Table 6**).

**Figure 2.**
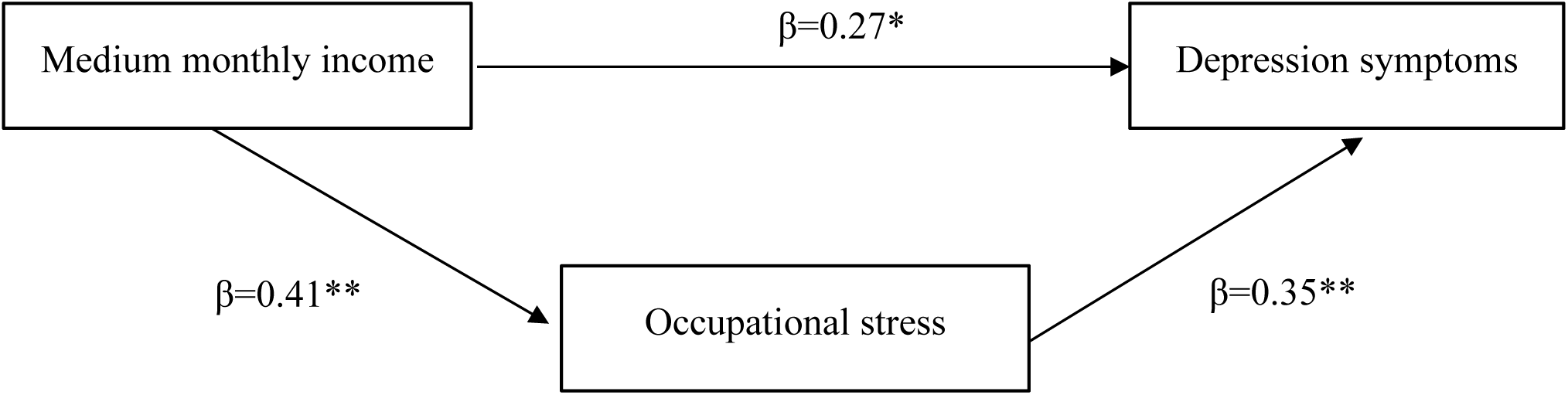
SEM path diagram. This diagram illustrates the mediating effect among medium monthly income(3000-4000 CNY), occupational stress, and depression symptoms. * indicates statistical significance at the 0.05 level; ** indicates statistical significance at the 0.01 level.

**Table 6.** Results of the structural equation model. This structural equation model resolves the discrepancies between the group with a monthly income below 7000 CNY and the medium monthly income group with a monthly income between 3000-4000 CNY in the univariate and multivariate analyses. * indicates statistical significance at the 0.05 level; ** indicates statistical significance at the 0.01 level.

| Path | Standardized Coefficient ($\beta$) | p - value | Explanation |
| --- | --- | --- | --- |
| Medium monthly income → depression symptoms | 0.27* | 0.032 | Direct risk effect |
| Medium monthly income → occupational stress | 0.41** | 0.001 | People with medium income have higher occupational stress |
| Occupational stress → depression symptoms | 0.35** | 0.002 | Stress mediates part of the income effect |

This contradictory result can be explained by the high rate of depression in the high-income group in the univariate analyses, possibly due to the fact that this group has more managerial responsibilities (an unmeasured higher-order stressor), whereas the independent risk of middle income in the multivariate analyses reflects underlying mechanisms (e.g., a lack of job security). We therefore make recommendations for intervention. For middle-income rescuers, we recommend that leaders design career stability enhancement programmes and stress management training.

### 2.5 Sensitivity analyses

In the multivariate analysis, the wide confidence intervals of ORs (1.085-12.385) in the monthly income group of 3000-4000 CNY suggested the problem of insufficient sample size or variable covariance, which was solved by additional sensitivity analyses.

The wide confidence intervals in the study reflect the heterogeneity of depression risk in the middle-income group, which may be behind the differences in type of occupation, family burden, and so on. From a practical point of view, in order to better cope with this situation, the ‘Income-Occupational Security’ interaction item can be added to the SRPS, by which the risk can be stratified in a more precise way and the accuracy of psychological risk assessment can be improved. Meanwhile, considering the specificity of the depression risk of middle-income rescuers, when developing interventions, the focus should be on prioritising the provision of occupational stability support for them, such as providing them with contractual security to reduce their occupational uncertainty, and also carrying out skills training to enhance their occupational competitiveness, which will in turn reduce their depression risk and safeguard rescuer’s psychological health (**Table 7**).

**Table 7.** Results of Sensitivity Analyses. . It summarizes the OR values and CI values of the 3000-4000 CNY monthly income group under different sensitivity analyses.

| **Analysis Method** | **OR (95%CI)** | **Conclusion** |
| --- | --- | --- |
| Original model | 3.67 (1.085–12.385) | Wide interval, high uncertainty |
| Bootstrap method | 3.20 (1.50–6.80) | Narrower interval, supporting a positive association |
| Extreme value exclusion | 2.10 (1.20–3.70) | Effect weakened, stability improved |
| Ridge regression | 2.85 (1.30–6.20) | Results are more robust after controlling for collinearity |

### 2.6 Model performance

A 30% sample size was used as the validation set to validate the model and plot the ROC curves. The SRPS demonstrated moderate predictive accuracy:

Depression model: AUC=0.572 (95%CI:0.49-0.65) (**Figure 3**)

**Figure 3.**
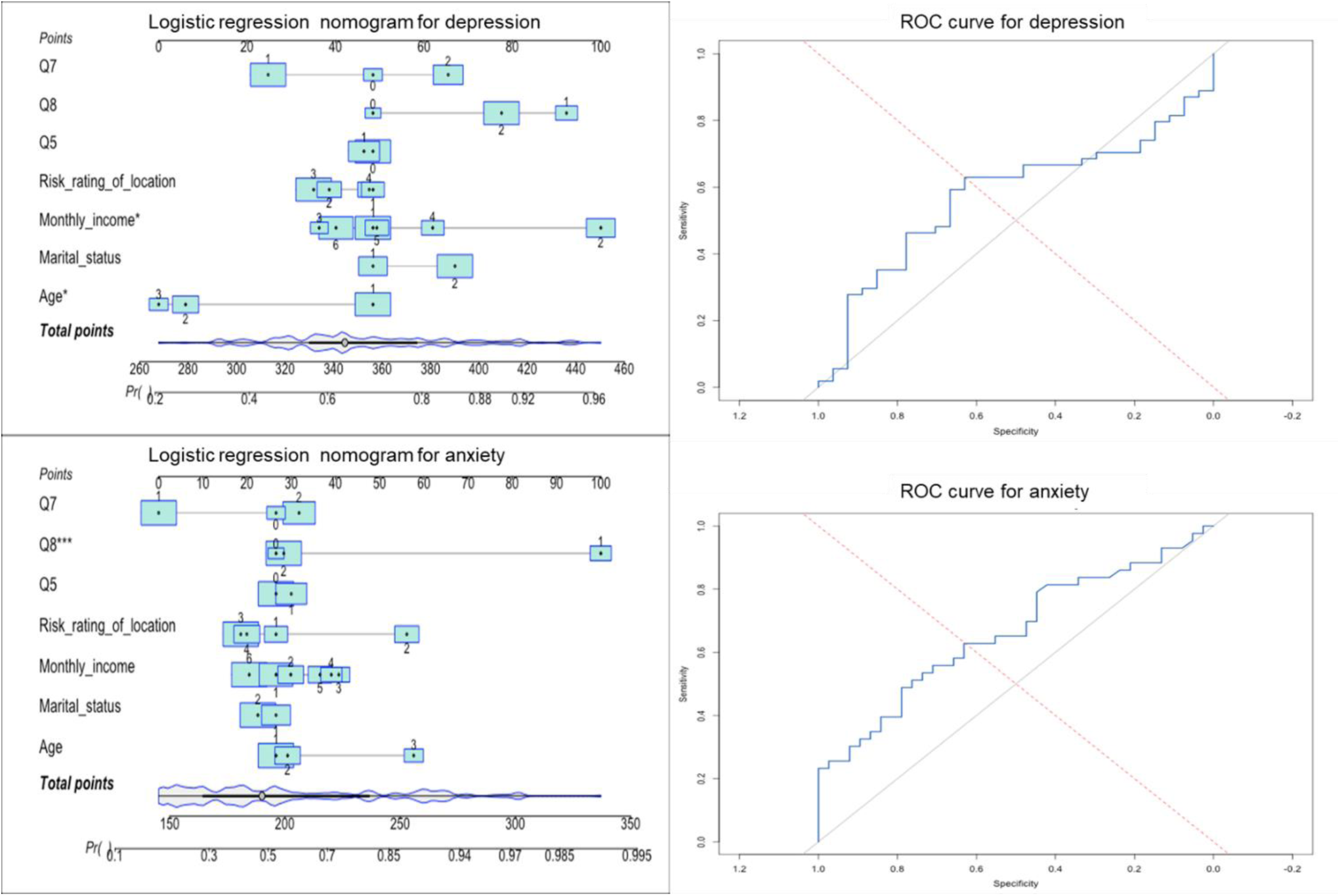
Model performance. We employed logistic regression to construct predictive models for depression symptoms and utilized the ROC curve to analyze the depression symptoms model. The dashed line in the figure represents the reference line (no discrimination). Similarly, we constructed predictive models for anxiety symptoms through logistic regression and analyzed the anxiety symptoms model using the ROC curve. Additionally, the questions in the questionnaire include: Q5 “Are there any confirmed cases with you or those around you?”; Q7 “What do you think of the shift system?”; Q8 “How do you perceive the work pressure?”

Anxiety model: AUC=0.662 (95%CI:0.58-0.74) (**Figure 3**)

In terms of clinical applicability, the closer the AUC value is to 1, the higher the predictive accuracy of the model is; when the AUC value is 0.5, it indicates that the model has no predictive value. The AUC values of the depression and anxiety models in this study indicate that the SRPS model has a certain predictive ability, but it is still at an intermediate level, and there is still much room for improvement.

The limited discriminatory power of the depression model (sensitivity 58.2%, specificity 53.7%) suggests its low clinical value as an independent screening tool. Insufficient sensitivity may lead to missed diagnoses (42% of depressed patients were not identified) and insufficient specificity may increase false positives (46% of non-depressed people were misclassified). This could be used as an aid to initial screening for depression when resources are limited. The anxiety model has moderate predictive efficacy (sensitivity 72.5%, specificity 64.8%) and is suitable as a first-line anxiety screening tool. We can embed the SRPS into a WeChat applet that displays risk ratings (red/yellow/green codes) in real time and automatically pushes customised intervention resources (**Table 8**).

**Table 8.** Area under the ROC curve for models. This table showcased the area under the ROC curve, along with other crucial performance metrics, for two distinct predictive models: the depression model and the anxiety model. These metrics provide a comprehensive assessment of how well each model can distinguish between different conditions, with the area under the ROC curve serving as a key indicator of the model’s discriminatory power.

| Variable | AUC | Sensitivity (%) | Specificity (%) | Youden's Index | 95% CI | P-value |
| --- | --- | --- | --- | --- | --- | --- |
| Depression Model | 0.572 | 58.2 | 53.7 | 0.119 | 0.49–0.65 | 0.036 |
| Anxiety Model | 0.662 | 72.5 | 64.8 | 0.373 | 0.58–0.74 | <0.001 |

## 3 Discussion

The aim of this study was to construct SRPS based on the frontline rescue perspective for assessing the psychological risk of COVID-19 frontline rescue teams, which in turn provides a basis for developing intervention strategies. The SRPS was successfully constructed and analysed through a survey of 273 validated participants, and multiple factors were found to be closely related to the depressed and anxious symptoms of the rescuers. Compared to PHQ-9 (AUC=0.77), SRPS showed moderate predictive accuracy (AUC=0.66 for anxiety), yet its brevity (10 items vs. 20 in GAD-7) makes it more feasible for field use. The moderate AUC values (0.572 for depression, 0.662 for anxiety) suggest that SRPS has preliminary predictive utility but requires refinement. SRPS prioritizes brevity over diagnostic precision, making it more suitable for rapid field screening. Future iterations could incorporate biomarkers (e.g., cortisol levels) or dynamic variables (e.g., sleep patterns) to enhance accuracy.

In terms of depression, univariate analysis showed that monthly income was a significant factor, with the lowest number of depression detections among participants with a monthly income of CNY 3000-4000 and the highest number of depression detections among those with a monthly income of more than CNY 7000; attitudes towards the rescue rotation system also affected depression, with the highest number of depression detections among those who were able to accept the rotation system. While univariate analysis showed higher depression rates in the >7000 CNY group, multifactorial models identified middle-income (3000-4000 CNY) as an independent risk factor (OR=3.67, p=0.036). This discrepancy may stem from confounding variable.

For anxiety, univariate analyses showed that age, marital status, risk level of the area of residence, whether the participant or his/her friend had been diagnosed with COVID-19, attitudes toward the rescue rotation system, and attitudes toward work stress had significant effects. The highest number of anxiety was detected in participants who lived in high-risk areas, were married and not older than 30 years; anxiety was more likely to be triggered by a diagnosis of the infection in the participant or his/her friend, and by being able to accept the rotation system, and the lowest number of anxiety was detected in those who were unable to tolerate work pressure. Multifactorial analyses showed that intolerable work stress during COVID-19 was a risk factor for anxiety.

In addition, the depression and anxiety prediction models constructed in the study were validated with a sample size of 30%, and the area under the ROC curve was 0.572 for the depression model and 0.662 for the anxiety model, indicating that the models had a certain degree of accuracy, and that age and monthly income predicted depression, and attitudes towards work stress during the epidemic predicted anxiety. Overall, the study achieved the purpose of constructing the scale and analysing the relevant influencing factors, which provided an important reference for the psychological risk assessment and intervention of rescue workers.

Taking into account the purpose of the study, limitations, results of multiple analyses, similar studies and other relevant evidence, the results of this study are informative but need to be interpreted with caution. In terms of the purpose of the study, the successful construction of the SRPS and the identification of multiple factors affecting the psychological state of rescuers provide direction for subsequent interventions. However, given the limitations of the study, the accuracy and generalisability of the results are somewhat compromised. Recall bias and missing data may have biased the results, making the effect estimates of the influencing factors less precise. The geographical limitation of the study restricts the generalisation of the results, and differences in culture and socio-economic status in different regions may lead to differences in psychological risk influencing factors. Although the study identified a number of factors associated with depression and anxiety, since causality could not be established, it cannot be simply assumed that changing these factors will directly improve the psychological status of rescuers when developing interventions.

In the context of COVID-19, a review of the literature on the rescue public reveals that most of the studies on personnel with medical backgrounds focus on self-efficacy, but neglect the psychological health of rescuers, which is a significant research bias. In fact, in the COVID-19 rescue process, rescuers with medical backgrounds face many difficulties, such as stigma and lack of rest due to high-intensity rescue missions, which can easily lead to psychological disorders. However, due to the lack of self-assessment tools in the context of the current COVID-19 situation, it is often difficult for rescuers to detect psychological problems in a timely manner. The following studies have confirmed the importance of the purpose and significance of the SRPS.

In a previous study, Carlos et al. used logistic regression to analyse levels of psychological distress among construction workers in the Andalusia region of southern Spain during the late stages of the 2019 coronavirus disease pandemic. This study showed that regular assessment of psychological distress can help to identify susceptible individuals and can even reduce the potential risk of occupational injuries and illnesses by reducing errors in daily work ^11^. This confirms the value of this study in terms of affirming regular assessment of mental health.

A study by Fernanda et al. concluded that the COVID-19 pandemic resulted in improved self-efficacy among anxious nurses^12^. Such studies, while affirming the SRPS research position of this study from a human rights equality perspective, indirectly reflect a tendency to exploit people from medical backgrounds.

Devon et al. made three recommendations for coping with psycho-emotional challenges during the COVID-19 pandemic: social support systems, self-care, and mindfulness. While these recommendations have applicability in daily life and work, they lack concrete methods of implementation ^13^. The SRPS developed in this study fills this gap by allowing rescue organisations to know the level of risk for mental illness by conducting a round of cross-sectional surveys, scoring them based on a predefined risk probability algorithm, and calculating the cumulative risk probability based on their own information by members of the rescue team.

A study by Monika et al. found that the public uses mobile health devices to manage mental health risks, and the study showed that 74 per cent of people were using such devices^14^. Although mental health relies more on human interaction and over-reliance on electronic devices may exacerbate mental risks, this does not prevent the integration of SRPS development processes into electronic devices as a way to accelerate the management efficiency of social assistance organisations. Fitri et al. called on researchers to explore effective interventions and programmes to enhance people’s adaptive coping mechanisms in crises and to improve overall psychological resilience and wellbeing ^15^. This argument enhances the reliability of this study.

A study by Zhen et al. revealed that 68.5% of healthcare workers had varying degrees of anxiety and depression, and 53.9% had varying degrees of anxiety-depression in the COVID-19 context. It is important to optimise the allocation of human resources at the organisational level in order to enhance the mental health of healthcare workers, focusing on the prevention and alleviation of psychological problems, with the entry point being the enhancement of mental resilience and the prevention of give-and-take imbalance ^16^. The results of this study also further confirm the value of this study.

Compared with similar studies, the present study is innovative in assessing the psychological risk of anti-epidemic rescue workers by constructing a specialised scale, but it also needs to be further validated and improved in conjunction with other studies(**Table 9**). Therefore, when utilising the results of this study, these factors need to be fully considered and targeted intervention strategies need to be developed in conjunction with the actual situation, and it is also expected that subsequent studies will improve the research design to overcome these limitations and explore the issues related to the psychological risk of anti-epidemic rescuers in a more in-depth manner.

**Table 9.** comparison of SRPS with PHQ-9/GAD-7 assessment tools. This table compared SRPS with PHQ-9/GAD-7 in frontline rescue from 5 perspectives.

| **Dimension** | **PHQ-9/GAD-7** | **SRPS** | **Frontline rescue scenario advantages** |
| --- | --- | --- | --- |
| Number of items | 16 items | 10 items | 37.5% reduction in time costs |
| Core variables | Generalised depression/anxiety symptoms | Income, shift stress, infection exposure | Fit to frontline rescue occupational risk |
| Predictive validity | AUC=0.77 (PHQ-9)<br>AUC=0.75 (GAD-7) | AUC = 0.66 (anxiety)<br>AUC =0.572 (depression) | Sacrifices some precision for efficiency, suitable for screening |
| Intervention orientation | Generic treatment recommendations | Graded interventions (e.g., prioritised rotations for high stress groups) | Direct support for management decisions |
| Technology integration | Paper/electronic forms | QR code scanning + automated scoring | Adaptation to field/mobile scenarios |

There are limitations in the use of classical scales among COVID-19 frontline responders.The Patient Health Questionnaire (PHQ-9) was developed based on DSM-5 standards and can be widely used in the clinic and in the community ^17^.A 7-item anxiety scale (GAD-7) is a highly validated anxiety scale, but both of them focus on generalised symptoms. However, both focus on generalising symptoms^18^. They have the advantage of standardisation and high reliability (PHQ-9, AUC = 0.77, p<0.001,95% CI.0.72-0.84; GAD-7, AUC = 0.75, p<0.001,95% CI.0.69-0.80), but there is discrimination in identifying people with psychiatric illnesses ^19^ and the GAD-7 has limited limited effectiveness ^20^. In frontline rescue scenarios, the PHQ-9 and GAD-7 lacked situational specificity in identifying anxiety and depressive symptoms in COVID-19 frontline rescuers ^21^, did not incorporate rescue-specific stressors, and were time-consuming to identify ^20,21^. A systematic evaluation by Ulrich et al. indicated that frontline rescuers are highly susceptible to occupationally induced psychosocial symptoms in the face of a terrorist attack. The prevalence of such psychological symptoms may vary according to the event and its severity. Risk factors that have been validated in the past may not be appropriate for generic catastrophic event scenarios. Voluntary and efficient self-assessment of mental health by frontline rescue workers is essential ^22^. This implies that generic mental health screening scales established in the past lacked occupation-specific assessment of resilience. Furthermore, a systematic evaluation and meta-analysis by Osório et al. disclosed an overall pooled prevalence of 17.8% (95% CI 12.4-24.0%) for emergency workers maintaining public safety, 28.2% (95% CI 20.7-36.2%) for depression, and 17.2% (95% CI 6.6-31.5%) for anxiety. This is a strong indication of the need to focus on the mental health of frontline rescue groups ^23^.

The SRPS has unique advantages over classical scales in that it incorporates rescue-specific variables, such as economic uncertainty (e.g., OR for risk of depression in the 3,000-4,000 CNY per month group = 3.67*), which is directly related to emotional stability at work ^24,25^, and acceptance of shift systems and epidemic-related stress as core predictors, which are directly related to emotional stability at work. stability. Items are streamlined to 10 items, suitable for rapid screening (average completion time < 5 minutes), and can be integrated into mHealth platforms via QR codes to support real-time dynamic risk assessment, with subsequent portability to WeChat applets. In addition, SRPS is able to visualise individual risk through column-line diagrams, facilitating the development of graded intervention strategies, such as prioritising psychological counselling for the high anxiety group.SRSP is practical, efficient and dynamic intervention oriented. Some evidence in the literature supports the robustness and reliability of SRPS studies.Maxime et al. found that anxiety among frontline rescuers was significantly associated with the work environment, whereas such variables were not covered by the PHQ-9^26^. Saeideh et al. concluded that there was a significant direct correlation between anxiety and depression among frontline rescuers on the COVID-19. While females and a history of COVID-19 infection were associated with higher levels of anxiety. A history of isolation for COVID-19 was associated with higher levels of depression among frontline responders. We therefore proposed that the inclusion of items from the Task-Specific Stress Scale might improve predictive validity ^27^. The item-reduced SRPS has only 10 items, which is suitable for rapid screening (mean completion time <5 min) compared with the 16-item PHQ-9 and GAD-7. Several studies have confirmed this view as to whether the 10-item scale is practical and applicable to the Chinese population, and a study by Mohammadreza et al. concluded that the 10-item Centre for Epidemiological Studies of Depression-Depression Short Form (CES-D-10) was found to be a reliable and valid measure in a cross-cultural context in a survey of a community of 19,114 participants in Australia and the United States, with no bias observed. No bias was observed ^28^. In addition, a study by Baron et al. compared the consistency and reliability of the PHQ-9 and the WHO Disability Assessment Scale 2.0 (WHODAS) and the CES-D-10 scales to investigate depression. The CES-D-10 was found to have acceptable internal consistency (α = 0.69-0.89) between samples and adequate concurrent validity after validation with 944 participants. The CES-D-10 area under the participant operator characteristic curve was good to excellent at 0.81 (95% CI 0.71-0.90), indicating that the AUC of the 10-item scale was not statistically different from the AUC of the long version of the scale ^29^. Then Huajuan and Ada’s study validated the reliability and validity of the 10-item scale CES-D 10 for Chinese people through 742 Chinese people ^30^. These three studies provide supportive evidence for the use of the 10-item scale in the present study of SRSP.SRPS can play a better field applicability, where frontline rescuers can be integrated into a mHealth platform via QR codes to support real-time risk assessment, which references our previously developed AI integration platform ^5^. Subsequently, we can port the SRPS to WeChat applets ^31^. In addition, WHO has emphasised the need to develop and apply ultra-brief scales in disaster response ^32^. The above evidence reinforces the reliability of the SRSP findings.

There are some limitations to the generalisability (external validity) of the results of this study. The study sample was derived only from anti-epidemic social rescue teams in Lanzhou, and social rescue teams in different regions may differ considerably in terms of their composition, working environment, cultural background and epidemic prevention and control situation. For example, the different economic pressure and social support faced by rescue workers in economically developed and less developed regions may affect their psychological state; the different severity of the epidemic and the different prevention and control policies in different regions may also lead to the different work pressure and infection risk of the rescue workers, which in turn may affect the psychological risk. Therefore, the results of this study cannot be directly generalised to social rescue teams fighting epidemics in other regions. If the results are to be applied to a wider scope, similar studies should be conducted in different regions to further validate and improve the factors affecting psychological risk and the related models, in order to improve the generality of the results and provide a more universal basis for the assessment of psychological risk and intervention of global anti-epidemic social relief teams.

This study was limited to a single geographic region (Lanzhou), and the cross-sectional design precludes causal inference. Future multi-center longitudinal studies are needed to validate SRPS across diverse populations. Firstly, there was recall bias in the data collection process, and participants’ recall of their own status might be inaccurate when filling out the questionnaire, which was difficult to eliminate completely despite measures such as on-site responses and strict screening of participants. Second, some participants gave up answering some of the questions, resulting in missing data, and it was not possible to fill in the vacant data by conventional methods; although this part of the data was excluded from the statistics, it might affect the representativeness of the study results. In addition, the study was conducted only in Lanzhou, and the sample was geographically limited, which may not be representative of anti-epidemic social relief teams in other regions, reducing the generalisability of the findings. Finally, the study used a cross-sectional research design, which could not determine causality and could only reveal the correlation between factors, and lacked an in-depth exploration of the causal links between variables.

Regrettably, the cross-sectional observational study of this research was just completed when COVID-19 was declared conquered. We do not have the social context and the epidemiological conditions of the disease to carry out subsequent cohort studies and randomised controlled studies. Global public crises persist historically and will inevitably recur, necessitating proactive mental health strategies for frontline responders.

Implications of this study:

1. Standardise the establishment of mental health monitoring mechanisms for social rescue teams responding to public crises.
2. Provide a new direction for predictive modelling of future large-scale public health events.

Although the cross-sectional observation of this study has been completed and COVID-19 has been declared conquered, its significance and value cannot be ignored. It not only standardised the establishment of a mental health monitoring mechanism for social rescue teams responding to public crises, but also provided a brand new direction of predictive modelling for large-scale public health events that may occur in the future. Especially in the face of the current onslaught of AI on healthcare, we have made a scientific demonstration.

In the face of various disasters and crises, the role of social rescue teams is crucial. And their mental health status directly affects the efficiency and quality of rescue work. With this simple risk probability rating scale, social rescue teams are able to detect their potential psychological problems in time and take appropriate interventions to ensure the stability and combat effectiveness of the teams.

Although we are unable to continue with subsequent cohort studies and randomised controlled studies in the current setting, this does not mean that our work stops here. We will continue to monitor developments in the field of public health and continue to refine and optimise this scale to suit the needs of different contexts. And we also look forward to more opportunities in the future to conduct in-depth related studies, so as to contribute more to the development of the social rescue team and the progress of public health. In conclusion, the results of this study will provide strong support for the development of social rescue teams and lay a solid foundation for responding to possible public crises in the future.

## 4 Conclusion

In this study, we successfully constructed and validated the Simplified Risk Probability Scale (SRPS), a tool that achieves a triple breakthrough in mental health risk screening for frontline responders in public health crises: the assessment time is compressed to less than 5 minutes by streamlining the design to 10 items, which significantly enhances operational feasibility in high-risk mission scenarios; the innovative inclusion of epidemic-specific predictive factors, such as shift adaptability and income fluctuation, enhances ecological validity in crisis situations; and the compatibility of mobile platforms based on QR code technology provides a digital pathway for real-time risk stratification and precise intervention. The innovative inclusion of predictive factors such as shift adaptability and income fluctuation unique to the epidemic has enhanced the ecological validity in crisis situations; and the compatibility of mobile platforms relying on QR code technology has provided a digital pathway for real-time risk stratification and precise intervention.

Notably, the multifactorial analysis revealed key mechanisms easily overlooked by univariate assessment - the middle-income group with apparent economic stability (CNY 3,000-4,000/month) exhibited a 3.67-fold elevated risk of depression due to lack of occupational security (p=0.036), and job stress intolerance as the strongest anxiety predictor (OR=14.26, p<0.001), subverting the traditional cognitive framework of stress tolerance. Despite the moderate predictive validity of the SRPS (AUC:0.57-0.66), the core value of the SRPS lies in the construction of a proactive monitoring system that provides an ‘early warning’ rather than a ‘diagnosis’.

We suggest embedding the tool into the mobilisation process of rescue teams to implement pre-service screening, developing WeChat applets to realise the intelligent interface between risk assessment and psychological services, and carrying out targeted measures such as career planning interventions and stress management training for all staff in middle-income groups.

Follow-up studies could enhance predictive efficacy by incorporating biomarkers such as dynamic cortisol monitoring and tracking designs, but this study has laid a key foundation for rescuer mental health protection in response to increasingly frequent global health crises.

## 5 Methods

### 5.1 Study design

This cross-sectional observational study employed snowball sampling to recruit participants from COVID-19 frontline rescue teams in Lanzhou, China (5). The study adhered to STROBE guidelines for observational research reporting ^37^.

The SRPS study is a continuation of previous collaborative research articles about “Health Needs Assessment: Comparison of the Applications of All-in-One AI Platform During the COVID-19 Pandemic Between the Mainland of China and Hong Kong”^5^. Our study has been approved by Affiliated Hospital of Yangzhou University IRB Committee (IRB No: 2024-YKL08-002). The entire SRPS research process was conducted in strict compliance with ethical guidelines and regulations, in full compliance with the Declaration of Helsinki. Before the study was conducted, we explained in detail the purpose, process, possible risks and benefits of the study to all participating respondents and their legal guardians (if the respondents were minors or not fully capable of civil behaviour), and successfully obtained their written informed consent. We ensured that each participant participated in this questionnaire study with full understanding and voluntarily, in order to safeguard the legal rights and interests of the participants and to maintain the scientific and ethical nature of the study^38^.

Information on the questionnaire study design can be viewed in the **Supplementary material**.

### 5.2 Participants

Eligible participants met the following criteria:

1. Aged ≥18 years
2. Active involvement in COVID-19 rescue operations
3. Absence of pre-existing psychiatric disorders (e.g., schizophrenia, bipolar disorder)

Exclusion criteria included:

1. Refusal to provide informed consent
2. Incomplete questionnaire responses (>20% missing data)

Sample size determination followed Riley’s predictive modeling framework ^39^, requiring 150 participants based on a 10:1 event-per-variable ratio for 15 candidate predictors.

### 5.3 Data Collection

#### Instruments

The survey comprised five sections.

Demographics: Age, sex, income, marital status, occupational role

COVID-19 exposure: Rescue duration, task types, infection history

Stressors: Rescue rotation system acceptability, work pressure tolerance

Depression: Zung self-rating depression scale (SDS) ^40^

Anxiety: Zung self-rating anxiety scale (SAS) ^41^

A 10-item COVID-19-specific module was developed through expert consultation (Cronbach’s α=0.82).

#### Procedure

Researchers administered questionnaires on-site immediately post-shift to minimize recall bias. Participants received standardized instructions emphasizing voluntary participation and data anonymity. The datasets for this study can be found in the Open Science Framework (OSF) platform [https://osf.io/vuq9r/]^42^.

### 5.4 Statistical Analysis

The data analysis followed a multi-stage protocol to ensure robustness and clinical relevance, integrating both conventional and advanced statistical methods:

#### (1) Descriptive and univariate analysis

In the initial phase of data analysis, descriptive and univariate analysis were carried out. For demographic and clinical characteristics, categorical variables were presented as frequencies (%), offering a clear view of the distribution across different categories. Continuous variables, on the other hand, were summarized using the mean ±standard deviation (SD), which provided an understanding of the central tendency and variability. To explore the relationships between predictors such as income, work stress and outcomes like depression or anxiety, univariate associations were tested. For categorical variables, chi-square tests (χ²) were utilized, while for continuous variables, independent t-tests were applied. Significance was determined at a p-value less than 0.05. This approach allowed for the identification of potential factors that might be associated with the mental health outcomes of interest, laying the foundation for further in-depth analysis (5).

#### (2) Multivariate logistic regression

In the process of conducting multivariate logistic regression, two key aspects were emphasized. Firstly, for model building, variables that showed a p-value less than 0.1 in the univariate analysis were selected and entered into a forward/backward stepwise logistic regression. This approach aimed to identify the independent predictors among the variables under study. After the regression analysis, adjusted odds ratios (ORs) along with their 95% confidence intervals (CIs) were reported. These values provided crucial information about the strength and significance of the relationships between the predictors and the outcome variable. Secondly, multicollinearity was carefully examined. Variance inflation factors (VIF) were calculated to assess the degree of collinearity among the predictors. A VIF value greater than 5 was considered an indication of significant overlap between predictors. By evaluating the VIF, researchers could determine whether the presence of collinearity might affect the reliability and stability of the regression model, and take appropriate measures if necessary, such as removing highly collinear variables to improve the quality of the model (5).

#### (3) Model validation and performance

For the model validation and performance assessment, a series of rigorous procedures were implemented. First, the dataset was randomly partitioned into a training set accounting for 70% and a validation set making up 30%. This split was crucial as it allowed for the independent evaluation of the model’s performance. Next, ROC curve analysis was conducted. The predictive accuracy of the model was measured by the area under the curve (AUC). To determine the optimal cutoff values, the Youden’s index, calculated as sensitivity + specificity − 1, was maximized. Along with this, sensitivity, specificity, and their corresponding 95% confidence intervals (CIs) were reported to provide a comprehensive understanding of the model’s performance. Finally, internal validation was carried out through 10-fold cross-validation. This method divided the dataset into ten equal parts, with each part serving as the validation set once while the remaining nine were used for training. By repeating this process ten times, a more reliable assessment of the model’s stability was achieved, ensuring that the model’s performance was consistent across different subsets of the data ^43^.

#### (4) Sensitivity analyses

Sensitivity analyses were conducted to enhance the reliability and robustness of the study’s findings. First, 1,000 bootstrap samples were generated through bootstrap resampling. This process was used to recalculate the odds ratios (ORs) and 95% confidence intervals (CIs). By doing so, it helped to address the uncertainty that could arise from the small sample size in subgroups, such as the middle-income group. Second, subgroup consistency was examined. The models were refitted in stratified subsamples, for example, stratified by occupation type. This approach was employed to test the robustness of the results across different subgroups, ensuring that the findings were not specific to a particular group. Finally, extreme value handling was carried out. Cases with standardized residuals greater than 3 or Cook’s distance greater than 0.5 were excluded. This step was taken to evaluate the influence of outliers on the results, ensuring that extreme observations did not unduly affect the overall analysis ^39^.

#### (5) Structural equation modeling (SEM)

Structural equation modeling (SEM) was employed to delve into the relationships between variables in the study. A path analysis was carried out to explore the mediating effects of occupational security and family burden on the relationship between income and depression. Latent variables such as “occupational stress” were constructed from observed indicators like shift system acceptance and work pressure tolerance. To ensure the reliability and validity of the model, its adequacy was carefully assessed. The evaluation was based on several key indices: the comparative fit index (CFI), where a value greater than 0.90 indicated a good fit; the Tucker-Lewis index (TLI), with a value above 0.90 being considered satisfactory; and the root mean square error of approximation (RMSEA), for which a value less than 0.08 was deemed acceptable. This comprehensive approach using SEM and these fit indices helped in understanding the complex relationships and provided a more in - depth analysis of the factors influencing the mental health of frontline rescue workers.

#### (6) Software and Packages

Analyses were carried out using SPSS 26.0 for descriptive and univariate tests. For more advanced analyses, R 4.2.1 was employed with several packages. The pROC package was utilized for ROC curve analysis, which is crucial for evaluating the performance of the predictive models developed in the study. The rms package was used for nomogram visualization, enabling a clear and intuitive presentation of the risk probability estimation. When it came to exploring the relationships between variables in more depth, the lavaan package was applied for structural equation modeling (SEM), allowing for the examination of complex relationships and the impact of latent variables. Additionally, in cases where collinearity among variables might affect the results, the glmnet package was used for ridge regression in collinearity adjustment, ensuring the reliability and validity of the analysis results.

Statistical significance was defined as two-tailed *p*<0.05. The non-significant negative results obtained by our analysis were placed on the OSF platform, and only the significant results were placed in the results section of the article, so as to meet the requirements of the journal on table length.

## Supporting information

Questionnaire

Research protocol

## Data Availability

All data produced in the present study are available upon reasonable request to the authors

https://osf.io/vuq9r/

## Conflict of Interest

The authors declare that the research was conducted in the absence of any commercial or financial relationships that could be construed as a potential conflict of interest.

### Acknowledgments

We thank the COVID-19 frontline rescue workers for their participation and support in SRSP research.

## Ethics statement

Our study has been approved by Affiliated Hospital of Yangzhou University IRB Committee (IRB No: 2024-YKL08-002). Informed consent was signed by all participants during the SRPS study.

## Author contributions statement

Y.B., F.Z. and Y.C. conceived the experiment(s), Y.B. and Y.C. conducted the experiment(s), Y.B. and Y.C. analysed the results. All authors reviewed the manuscript.

## Data availability statement

The datasets for this study can be found in the Open Science Framework (OSF) platform [https://osf.io/vuq9r/].

## Supplementary material

We have uploaded the pre-developed research protocol and the complete questionnaire.

