## Supplementary material for "Health Needs Assessment: Development and Validation of a Simplified Risk Probability Scale (SRPS) for Mental Health Screening in COVID-19 Frontline Rescue Teams": Research protocol

### 1 Introduction

The COVID-19 pandemic has profoundly affected people's daily life since December 2019. The virus has progressed into pulmonary fibrosis rapidly and caused hypoxic mortality by pulmonary fibrosis [1]. Since the high mutation rate makes it challenging for common antiviral drugs to exert clinical therapeutic value, residents must restrict their outdoor activities for self-protection [1]. However, during the COVID-19 epidemic, China tried the herd immunity barrier to eliminate the pandemic [2]. The HK government dispatched vanguards to block the spread of COVID-19, including input from rescue teams composed of non-medical professionals such as security guards and volunteers [3].

Frontline rescue workers were more likely to become a high-risk group for depression and anxiety due to the long-term coping with much work for patients and the inability to keep contact with close family members [4,5]. During the pandemic, the detection rates of anxiety and depression in Hong Kong's anti-epidemic rescue teams reached 19.8% and 27.7% [6]. Infection with the virus, emotional loneliness, separation from family members, inconvenience, and age were considered significant factors for depression and anxiety [4,7–9]. However, it is difficult for the rescue vanguard to use these research results for self-examination when carrying out rescue missions [6]. Establishing a new risk prediction model based on previously reported critical factors related to psychological disorders is essential in the post-pandemic period, as making a simple risk score table and accumulating the scores according to their conditions [10]. In response, it's important to note that developing such a model would require rigorous research and validation to ensure its accuracy and effectiveness. Mental health professionals and researchers must collaborate closely with rescue organisations to gather data, identify the most relevant factors, and create a practical and reliable tool for use in the field.

The present study endeavours to enhance the precision of mental health assessment of anti-epidemic rescuers. Therefore, the risk probability score table can predict their psychological conditions and develop more informed intervention strategies. Unlike PHQ-9 (AUC=0.85) and GAD-7 (AUC=0.89), which require 10–15 minutes for completion, SRPS prioritizes brevity (10 items) and context-specific factors (e.g., rescue rotation system, income instability), enabling real-time screening during high-pressure missions.

### 2 Methods

#### 2.1 Study design

The present SRPS study employed a cross-sectional observational research design.We planned at that time to complete the SRPS study using questionnaires. The STROBE check terms developed by von Elm et al. [11] were followed throughout the SRPS report.

#### 2.2 Setting

The SRPS study started on 1 October 2022, and the period for recruiting participants to take part in the questionnaire was set at 2 months. This time period may change as the SRPS study faces difficulties, and any specific changes will be disclosed in the study results. The city where the questionnaire was administered was set to be Lanzhou, China.The exposure factors of interest to participants in the SRPS study were the 10 questions answered by the participants and the participants' state of anxiety and depression (Appendix).The main purpose of the SRPS study was to establish a self-assessment methodology, however, the COVID-19 social rescuers may only experience anxiety and depression when they are currently on a rescue mission, so setting up a follow-up was unnecessary. Data was collected and collated by recovering, reading, and coding all participant responses completed on the questionnaire placed into Excel.

### 2.3 Participants

During the execution of the cross-sectional observational study using questionnaires, in order to ensure a minimum sample size to achieve statistical power [12], the snowballing statistical principle [5] was employed to advertise the study and liaise with COVID-19 social rescue organisations. The snowballing execution essentially involved first contacting a social rescue team that was on a mission to rescue COVID-19 and subsequently convincing them to participate in the study. News of the content of the SRPS study was then diffused to other social rescue teams through members of this social rescue team. Secondly, we then publish the news of the SRPS study on the Internet and in the self-publishing media, and those social rescue organisations that are interested in the results of our anticipated research will contact us on their own initiative. Our feedback to participants is to share our research results unconditionally so that these participating social rescue teams will be the first to apply our SRPS.

Inclusion criteria for participants: (1) aged 18 years or older; (2) being involved or having been involved in COVID-19 social rescue; (3) having the basic ability to participate in the SRPS study, e.g., reading the questionnaire in paper form or electronically, and having no language communication barriers. Exclusion criteria for participants: (1) refused to sign the informed consent form in the first part of the questionnaire; (2) had doubts about the SRPS study but did not contact the study leader in a timely manner; and (3) the participants had psychiatric disorders such as schizophrenia, bi-directional depression, and other mental illnesses that affected the results of the questionnaire assessment.

### 2.4 Variables

We produced the questionnaire in October 2022 at that time. This questionnaire consisted of five parts (**Appendix**). Items were refined based on expert feedback to enhance content validity. Data collection followed a two-phase snowball sampling protocol, with on-site administration to minimize recall bias. The first section described the content and purpose of the SRPS study and the informed consent form. Social assistance teams who were given the questionnaire were asked to sign the informed consent form by hand if they were interested in the SRPS study, otherwise they were deemed to have refused to sign the informed consent form. The second section contained mainly demographic information and questions related to current COVID-19 rescue efforts. The third section was designed as a multiple choice question with the intention of asking COVID-19 social rescue members where their stress comes from during the rescue process. The fourth section was the Depression Self-Rating Scale, which referenced the Depression Self-Rating Scale developed by Zung in 1965 [13]. The fifth section was the Anxiety Self-Assessment Scale and this referenced the Anxiety Self-Assessment Scale developed by Zung in 1971 [14]. Throughout the completion of the questionnaire, we allowed full response, partial response, or abstention. This was based on our adherence to the ethical guidelines for medical research at Sindal. Participants were allowed to choose not to answer certain questions when they perceived that they would feel uncomfortable with disclosing their privacy, and we had to fully respect the participant's choice. This study was conducted in accordance with the 1964 Declaration of Helsinki and its subsequent amendments or similar ethical standards.

The exposures of ultimate interest for the SRPS study are anxiety and depression. The definitions of anxiety and depression follow the requirements of the DSM-5 [15]. As it was not always possible to send a psychiatrist or psychologist to confirm the diagnosis of illness in the SRSP study, we used the SAS [14] and SDS [13] to assess the participants. Positive results assessed here indicate only anxiety symptoms and depressive symptoms, and the severity of the assessment only represents the severity of the symptoms or the probability that anxiety or depression may occur, and does not imply that the participant has a confirmed diagnosis of anxiety or depression.

Potential confounders affecting exposure in the SRPS study were demographic information and 10 questions answered by participants (**Appendix**).

In addition, the definition of COVID-19 may interfere with the accuracy of participants' completion of the questionnaire, so for the definition of COVID-19, we referred to the local treatment guidelines and expert consensus in Lanzhou: the diagnosis of infection with COVID-19 was confirmed by the presence of COVID-19 nucleic acid positivity in human specimens collected from nasal swabs, pharyngeal swabs, and venous blood after testing . In our study, there was no deliberate emphasis on COVID-19 pneumonia or pulmonary symptoms. This was because confirmation of COVID-19 pneumonia or pulmonary symptoms required participants to go to the hospital for radiological testing, whereas in the prevailing environment of the COVID-19 pandemic, nasal swabs, pharyngeal swabs, and venous blood specimens were collected on a daily basis, and each person's e-health code contained information on COVID-19 nucleic acid testing, which was more in line with ethical requirements for medical research than radiological testing. This is more in line with the ethical requirements of medical research than radiological testing. It is worth noting that whenever a person is able to go out on a COVID-19 rescue mission, the COVID-19 nucleic acid test results of these social rescue workers are negative, i.e. they are not infected with COVID-19.

### 2.5 Data measurement

The data were derived from questionnaires that were read, collated and coded for recovery by the study sponsor, Yan Bo. In it, demographic information and predefined 10 questions participants could answer directly or make options (Appendix).

Measures of anxiety and depression were converted according to scoring rules developed by the SAS [14] and SDS [13]. A score of less than 50 is considered ‘asymptomatic’, while a score of less than 50 is considered ‘symptomatic’. In other words, a participant's ‘symptomatic’ outcome is considered to be the presence of depression or anxiety in the SRSP study. Note that the presence of depression or anxiety is not the same as a diagnosis of depression or anxiety. The ‘absence of symptoms’ does not necessarily mean that a diagnosis of depression or anxiety will not be made. This requires a rigorous assessment by a psychiatrist or psychologist. Studying only anxious and depressive symptoms was done to better complete this SRPS study during the COVID-19 pandemic.

### 2.6 Bias

The bias present in the SRSP study comes from two main sources: bias in participants' recall of their own state when completing the questionnaire and abandonment of answering some of the questions. For questionnaires that were abandoned to answer some of the questions, we dropped this part of the data in the final statistics. We were unable to fill in the data values that were left vacant due to the abandonment of responses through conventional mean and interpolation methods. The results of the study will disclose the ratio of partial to full responses. To address our own recall bias, we will travel to the site of the COVID-19 social rescue team's mission, ask the social rescuers to respond on-site, and collect the questionnaires on-site. In addition, we will also assess the social social rescuers prior to their participation in the study, i.e., the previously mentioned inclusion and exclusion criteria. In addition, in order to reduce statistical bias, we used both univariate and multivariate analyses of the factors affecting anxiety and depression, and instead of using separate P-values, we used P-values to jointly test for the co-occurrence of effect sizes in the final report of the results[16].

To minimize recall bias, all questionnaires were administered on-site during rescue missions. Participants completed the survey immediately after their shifts, with researchers present to clarify any ambiguities. This real-time data collection strategy ensured higher accuracy in self-reported psychological states.

While snowball sampling enhanced accessibility during pandemic restrictions, it may have introduced selection bias (e.g., overrepresentation of participants from similar organizational backgrounds).

### 2.7 Study size

The minimum theoretical sample size for assessing predictive model types in the SRSP study was assessed using a method written by Riley [12]. It is expected that a predictive model will be built using 15 variables, which would then require 150 samples according to the 10-fold assessment method according to Riley.

### 2.8 Quantitative variables

The quantitative variables in the SRPS study were demographic information and responses to 10 questions (**Appendix**). We plan to use chi-square tests and binary logistic regression models for the quantitative variables.

### 2.9 Statistical methods

In the SRPS study, set statistical p-values are all considered significant at $P<0.05$ for bilateral tests. Significant p-values need to be marked with ‘*’ in the data table. In order to facilitate the understanding of the statistical process and the replicability of the study, a set of statistical steps was established:

(1) All collected questionnaire data were descriptively analysed to observe the overall characteristics of depression and anxiety in the new crown rescue team in Lanzhou.

(2) In order to investigate the correlation that exists between demographic informatics and depression and anxiety in the New Crown Rescue Team, a chi-square one-way stratified analysis was conducted.

(3) In order to investigate the existence of associations between facing the 10 questions of the self-administered New Crown questionnaire and depression and anxiety in the New Crown rescue teams, chi-square one-way stratified analyses were conducted.

(4) Since it was already known through univariate analyses which variables individually correlate with depression or anxiety, depression or anxiety is usually influenced by multiple factors

simultaneously. Therefore, we need to use binary logistic regression model to test which variables are correlated to anxiety or depression at the same time. The model can be adjusted to the best during this period using the overall inclusion method, forward and backward methods, etc.

(5) We attempted to develop models that predicted depression and anxiety, thus better allowing the Lanzhou New Crown Rescue Team to conduct self-psychological assessments. We incorporated the previously significant single and multifactorial variables into the predictive model.

(6) The performance of the depression and anxiety models was assessed and ROC curves were plotted.

(7) Visualise the established predictive models using column-line plots.

In the statistical process of the above 7 steps, categorical variables were compared using chi-square test or Fisher's exact test. Continuous variables were expressed as median, minimum, or maximum values, and these statistical schemes followed those used in a previous survey of both mainland China and Hong Kong [5]. Model validation utilized 10-fold cross-validation, with ROC curve thresholds set at Youden's index to optimize sensitivity and specificity. For assessing model performance using ROC curves as well as column-line graphs to visualise the predictive model refer to Wang et al. We placed the significant one-way and significant multifactorial distributions of depression and anxiety described above into multifactorial logistic regression for modelling. We used 70% of the sample size as a training set to construct the predictive model and plot the column line graphs [17].
